# Mucosal immunity against SARS-CoV-2 variants of concern including Omicron following vaccination

**DOI:** 10.1101/2022.01.26.22269659

**Authors:** Jinyi Tang, Cong Zeng, Thomas M. Cox, Chaofan Li, Young Min Son, In Su Cheon, Yue Wu, Supriya Behl, Justin J. Taylor, Rana Chakaraborty, Aaron J. Johnson, Dante N Shiavo, James P. Utz, Janani S. Reisenauer, David E. Midthun, John J. Mullon, Eric S. Edell, Mohamad G. Alameh, Larry Borish, Mark H. Kaplan, Drew Weissman, Ryan Kern, Haitao Hu, Robert Vassallo, Shan-Lu Liu, Jie Sun

**Author notes:** These authors contribute equally (Jinyi Tang, Cong Zeng). These authors contribute equally (Shan-Lu Liu, Jie Sun).

## Abstract

SARS-CoV-2 mRNA vaccination induces robust humoral and cellular immunity in the circulation; however, it is currently unknown whether it elicits effective immune responses in the respiratory tract, particularly against variants of concern (VOCs), including Omicron. We compared the SARS-CoV-2 S-specific total and neutralizing antibody (Ab) responses, and B and T cell immunity, in the bronchoalveolar lavage fluid (BAL) and blood of COVID-19 vaccinated individuals and hospitalized patients. Vaccinated individuals had significantly lower levels of neutralizing Ab against D614G, Delta and Omicron in the BAL compared to COVID-19 convalescents, despite robust S-specific Ab responses in the blood. Further, mRNA vaccination induced significant circulating S-specific B and T cell immunity, but in contrast to COVID-19 convalescents, these responses were absent in the BAL of vaccinated individuals. Using an animal immunization model, we demonstrate that systemic mRNA vaccination alone induced weak respiratory mucosal neutralizing Ab responses, especially against SARS-CoV-2 Omicron; however, a combination of systemic mRNA vaccination plus mucosal adenovirus-S immunization induced strong neutralizing Ab response, not only against the ancestral virus but also the Omicron variant. Together, our study supports the contention that the current COVID-19 vaccines are highly effective against severe disease development, likely through recruiting circulating B and T cell responses during re-infection, but offer limited protection against breakthrough infection, especially by Omicron. Hence, mucosal booster vaccination is needed to establish robust sterilizing immunity in the respiratory tract against SARS-CoV-2, including infection by Omicron and future variants.

## Introduction

The ongoing COVID-19 pandemic is a global public health crisis, and vaccination is considered the key to ending the pandemic (*1, 2*). It is well recognized that current SARS-CoV-2 vaccines, particularly mRNA-based vaccination, can induce robust humoral and cellular immunity and prevent severe disease caused by SARS-CoV-2 (*3*); however, protection against non-symptomatic to mild infection and transmission by mRNA vaccination is relatively moderate (*4*), and the reasons for this are poorly defined. Notably, most of the previous studies were conducted using blood to determine circulating antibodies and B and T cell immunity following vaccination (*5*), and characterization of respiratory mucosal immunity is lacking, which is essential for understanding vaccine- and natural infection-mediated protection against SARS-CoV-2. Additionally, the newest SARS-CoV-2 variant of concern (Omicron), has been shown to easily escape both vaccine and infection-elicited Ab neutralization in the blood (*6-12*). However, it is currently unclear whether efficient mucosal neutralizing Ab responses can be induced by vaccination, and/or natural infection, and to what extent this could protect against SARS-CoV-2 infection. The current study was designed to address these critical questions, and our results demonstrated that robust mucosal immunity can be elicited in the lung by natural infection and mRNA vaccination plus adenovirus-mediated vaccination, but not by the mRNA vaccination alone, and that the induced mucosal immunity is essential for protection against infection by SARS-CoV-2, including the Omicron variant.

## Results

### Characterization of respiratory mucosal antibody responses following vaccination or natural infection

To determine the humoral and cellular immune responses following COVID-19 vaccination, we collected blood and BAL samples from a cohort of COVID-19-vaccinated individuals (Fig.1A). Most of these individuals had received two doses of mRNA vaccination, with 3 individuals receiving the third booster and one having the J&J vaccine. The vaccine type, timing of collection, age, and sex information are included in Extended Table S1. We compared the vaccine-induced respiratory and circulating Ab, as well as cellular immune responses, to those of hospitalized COVID-19 convalescents that we have previously recruited (*13*). We first performed enzyme-linked immunoassay (ELISA) to determine and compared the SARS-CoV-2 S1 or receptor binding domain (RBD)-specific IgG, IgA and IgM levels in unvaccinated control (non-SARS-CoV-2 infected), vaccinated, and convalescent groups in the plasma. Similar to what was shown before (*3, 14*), COVID-19 vaccination induced robust S1 or RBD-specific plasma IgG at levels comparable to severe cases of natural infection (Fig. 1B and Fig. S1A). The S1 or RBD-specific IgG levels in the BAL were also comparable between COVID-19-vaccinated and convalescent groups (Fig. 1C and Fig. S1B). Of note, COVID-19 vaccination failed to induce significant levels of systemic S1 or RBD-specific IgA, while COVID-19 convalescents exhibited moderate but detectable S1-specific IgA responses in the blood (Fig. 1D and Fig. S1C). Importantly, prior severe SARS-CoV-2 infection provoked significant levels of S1 or RBD-specific IgA in the respiratory mucosa, which was not the case for COVID-19 vaccination (Fig. 1E and Fig. S1D). We also examined IgM in the blood and BAL, and observed that, while detectable levels of IgM were present in the circulation of both COVID-19-vaccination group and prior infection cases, only prior infection elicited significantly elevated IgM responses in the BAL (Fig. S1E-H). Together, these results revealed that, in contrast to natural infection, COVID-19 vaccination does not provoke robust IgA or IgM responses in the respiratory tract.

**Fig. 1.**
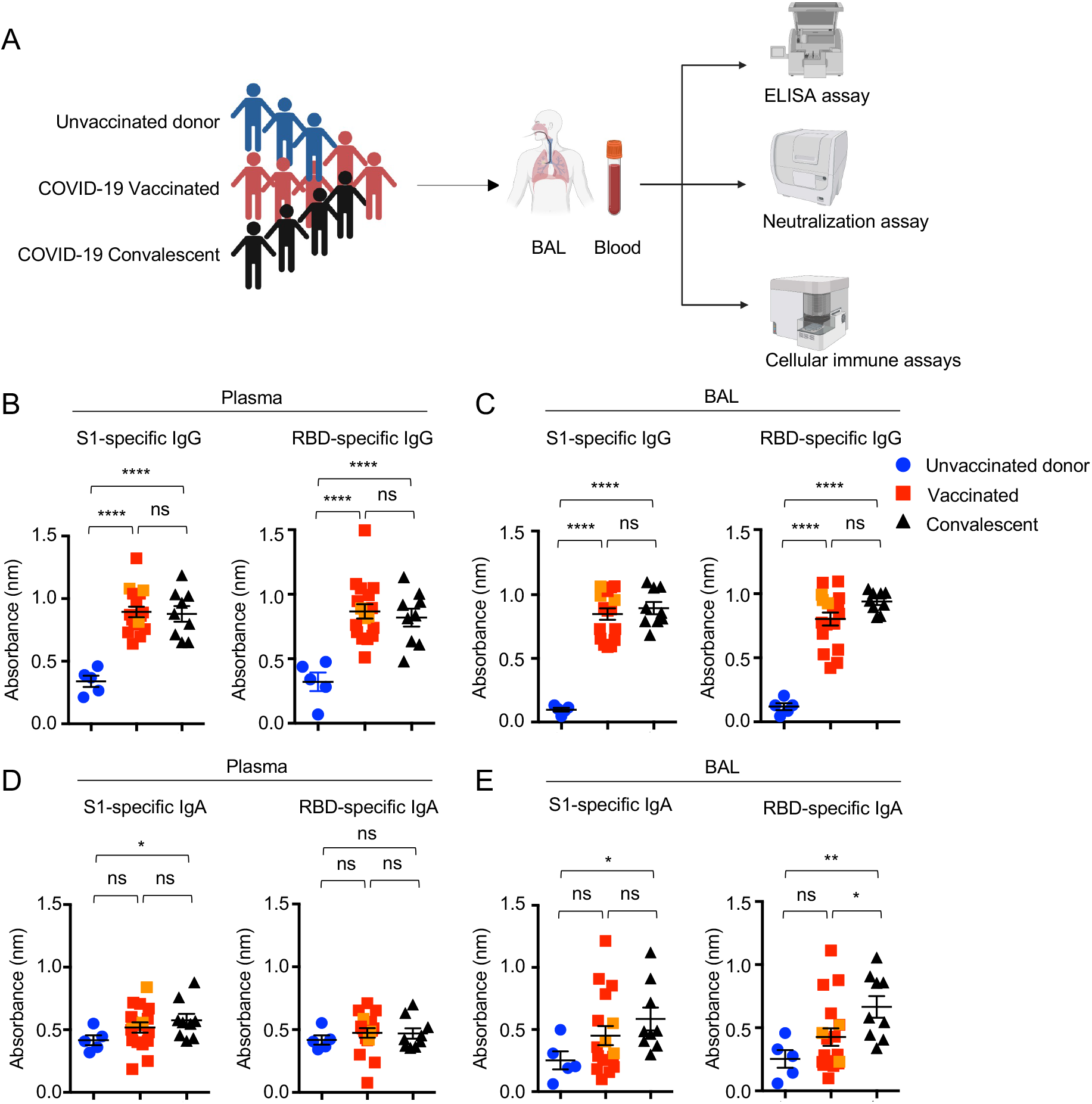
Systemic and respiratory antibody responses in COVID-19 convalescents and vaccinated individuals. **(A)** Schematic of recruited cohorts (n=5 for unvaccinated donor, n=19 for vaccinated, and n=10 for COVID-19 hospitalized convalescent) and experimental procedures. Figures were created with BioRender. **B to E**, Levels of SARS-CoV-2 S1 or RBD binding IgG **(B and C)** or IgA **(D and E)** in plasma and bronchoalveolar fluid (BAL) of unvaccinated donors, COVID-19 vaccinated or convalescents. Three individuals who received the booster (BNT162b2 or mRNA-1273) were indicated as orange in the vaccinated group. Enrolled donors’ demographics were provided in Extended Data Table 1. and a previous publication (*13*). Data are means ± SEM. Statistical differences were determined by one-way ANOVA and p values were indicated by ns, not significant (P > 0.05), * (p < 0.05), ** (p < 0.01), and **** (p < 0.0001).

### Mucosal Ab neutralizing activity against VOCs

The humoral protection against SARS-CoV-2 infection relies on the induction of robust neutralizing Ab (*15-17*). We thus examined the plasma neutralizing Ab activity against SARS-CoV-2 D614G, Delta and Omicron spike-pseudotyped lentiviruses (*18*) (Fig. 2 A-C and Fig. S2 A-C). While COVID-19 vaccinated and convalescent individuals exhibited comparable high levels of circulating neutralizing Ab responses against all viruses, the Delta and Omicron variants exhibited more than 2- and 10-fold decrease in neutralization titer (NT_50_), respectively, compared to D614G (Fig. 2 A-C), consistent with recent results showing that VOCs, especially Omicron, have significant immune evasion capability (Fig. 2C) (*6-12, 19-21*).

**Fig. 2.**
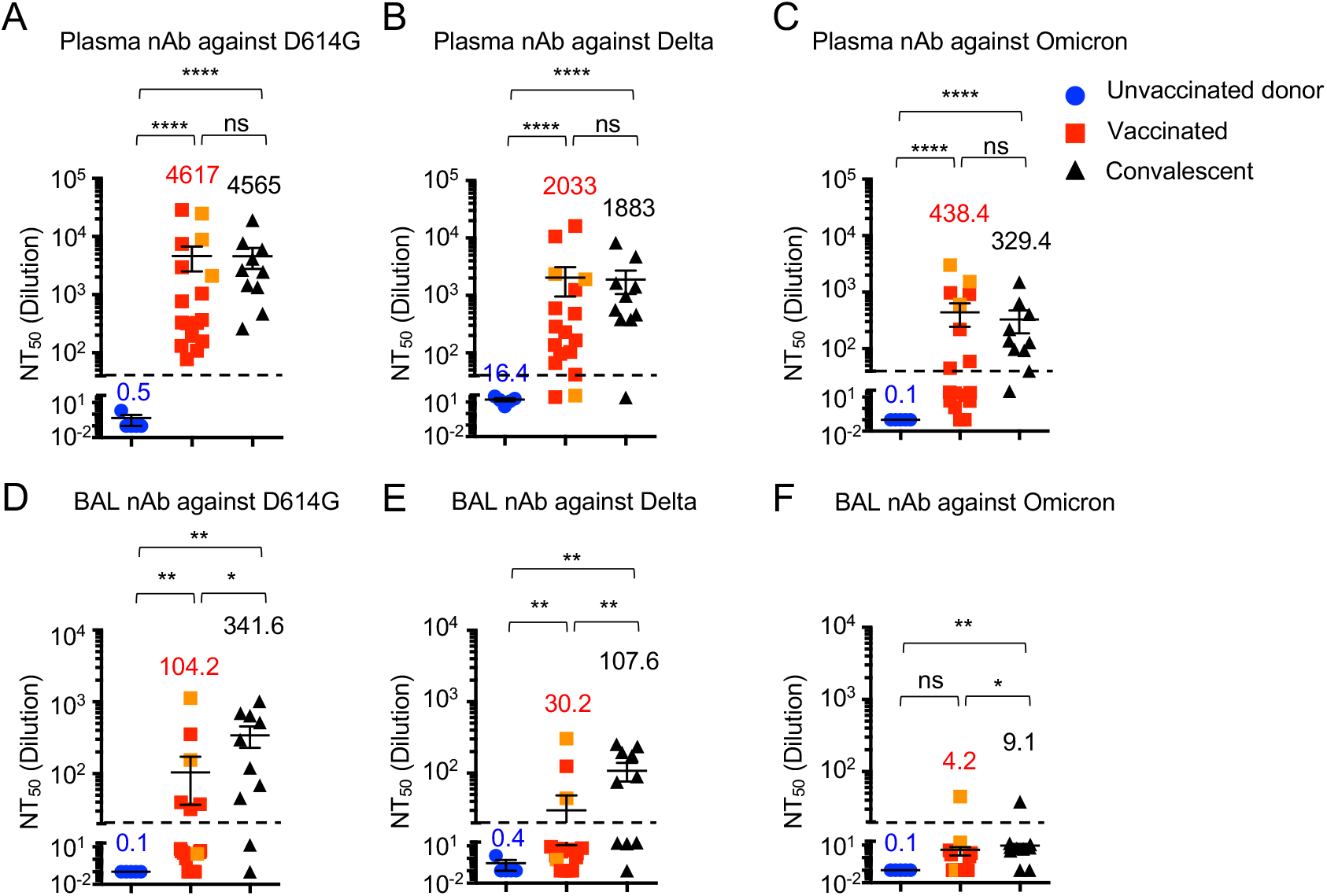
COVID-19-vaccinated individuals exhibit lower respiratory neutralizing antibody responses compared to convalescents. Plasma and BAL neutralizing activity in unvaccinated donors, vaccinated and convalescent individuals. (**A to C)** Neutralizing antibody titers (NT_50_) in plasma against SARS-CoV-2 S D614G (A) Delta (B) and Omicron (C) pseudotyped virus in unvaccinated donors (n=5), vaccinated (n=19) and convalescent (n=10) individuals. HEK293T-ACE2 cells were used as targeted cells for infection. **(D to F)** Neutralizing antibody titers (NT_50_) in BAL against SARS-CoV-2 S D614G (D), Delta (E) and Omicron (F) pseudotyped virus in unvaccinated donor, vaccinated and convalescent individuals. Three individuals receiving a booster shot (BNT162b2 or mRNA-1273) were indicated as orange in the vaccinated group. nAb, neutralizing antibody. Data are means ± SEM. Statistical differences were determined by one-way ANOVA and p values were indicated by * (p < 0.05), ** (p < 0.01), and **** (p < 0.0001).

We next compared neutralizing Ab responses in BALs of COVID-19-vaccinated and convalescent groups along with healthy controls. Despite the overall lower neutralizing antibody levels in BAL compared to that in the blood, the convalescent group showed ∼3-fold higher neutralizing Ab activity than the vaccinated group, especially for the ancestral D614G (p < 0.05) and the Delta variant (p < 0.01) (Fig. 2D-E). The titers for the Omicron variants were mostly below the level of detection (Fig. 2F), reflecting the stronger escape of Omicron from neutralizing antibodies. Of note, one out of three who had received a booster vaccination exhibited above-the-threshold yet low level of neutralization activity against Omicron (Fig. 2F), suggesting that booster vaccine may offer some, but limited, level of protection (Fig. 2F). Overall, these results indicate that natural infection elicits stronger humoral immunity in mucosal surface compared to mRNA vaccination, and better vaccine strategies are needed to offer protections against VOC, especially the Omicron variant.

### Mucosal cellular immunity is induced in COVID-19 convalescents, but not in mRNA vaccinees

Although memory T and B cells do not confer sterilizing immunity, they are important in constraining viral dissemination and protecting against severe diseases once a virus breaches neutralizing humoral immunity (*15, 22-25*). Both circulating and tissue-resident memory T and B cells are believed to provide disease protection against severe respiratory viral infection (*22, 26-28*). We therefore examined systemic and tissue residing memory T and B cell responses following mRNA vaccination or natural infection. Compared to unvaccinated controls, vaccinated individuals had higher RBD-specific B cells in the blood (Fig. S3 A-D). Notably, RBD-specific B cells were markedly lower in BAL compared to those of PBMCs (Fig. 3A and Fig. S3E). As reported before (*22, 29-31*), vaccination induced notable S-specific TNF or IFN-γ producing CD8^+^ or CD4^+^ T cells in the circulation, but failed to elicit strong S-specific cytokine-producing CD8^+^ or CD4^+^ T cell responses in the BAL (Fig. 3B, C and Fig. S4). In contrast, convalescent BAL exhibited much higher RBD-specific B cells compared to the paired blood samples (Fig. 3D), suggesting that vaccination does not induce tissue-residing memory B cell responses as effectively as natural infection. Further, BAL from COVID-19 convalescents had higher cytokine-producing CD8^+^ and CD4 T^+^ cells than those of blood (Fig. 3E, F and (*13*)). Within the total CD8^+^ or CD4^+^ T cell compartments, the levels of most memory T cell subsets in the blood and/or BAL were quite similar between unvaccinated or vaccinated individuals, except the blood TCM population (Fig. S4). Thus, unlike SARS-CoV-2 natural infection, mRNA vaccination does not appear to induce significant SARS-CoV-2 specific B and T cell memory in the respiratory mucosa in contrast to that in the blood.

**Fig. 3.**
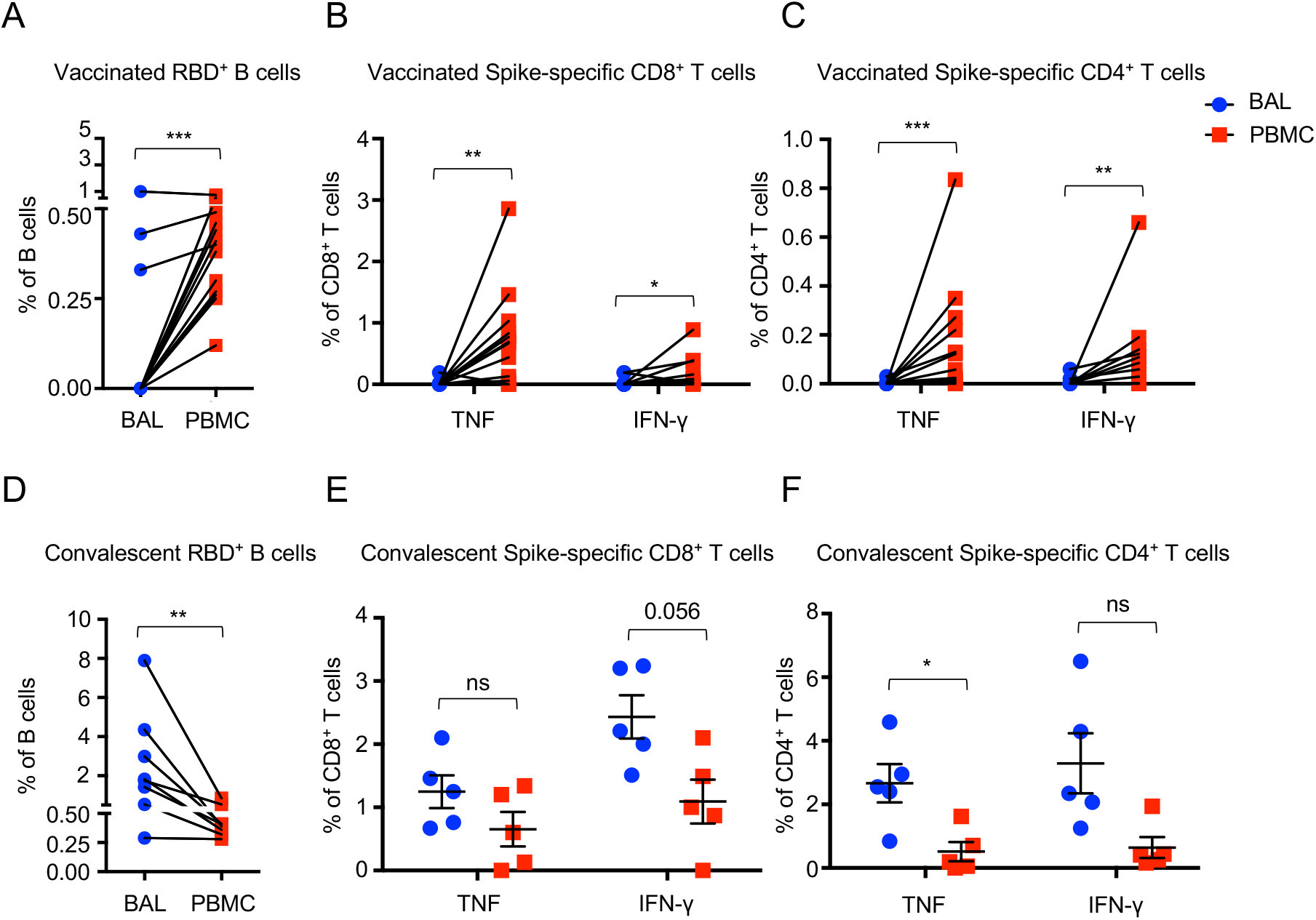
COVID-19-vaccinated individuals exhibit systemic cellular immunity not evident in the respiratory tract. **(A)** Frequency of SARS-CoV-2 RBD-specific B cells in the blood (PBMC) and the BAL of vaccinated. **(B and C)** Frequencies of TNF- and IFN-γ-producing CD8^+^ (B) or CD4^+^ (C) T cells in the blood (PBMC) and the BAL of vaccinated after S peptide pools stimulation. **(D)** Frequency of SARS-CoV-2 RBD-specific B cells in the blood (PBMC) and the BAL of convalescent individuals. **(E and F)** Frequencies of TNF- and IFN-γ-producing CD8^+^ (E) or CD4^+^ (F) T cells in the blood (PBMC) and the BAL of convalescents after S peptide pools stimulation. Data are means ± SEM. Statistical differences were determined by paired t-test in a-d and independent t-test in e-f. P values were indicated by ns, not significant (P > 0.05), ** (p < 0.01), and *** (p < 0.001).

### mRNA plus mucosal Ad5-S booster induces strong neutralizing immunity against Omicron

Given the suboptimal mucosal immunity induced by the current mRNA vaccination, we used an animal model to identify potential strategies that promote and/or amplify mucosal humoral and cellular immunity after mRNA vaccination. To this end, we immunized wildtype mice with PBS, two doses of mRNA-encoding codon-optimized S (mRNA-S), three doses of mRNA-S, two doses of mRNA-S plus an intranasal immunization of S protein trimer with adjuvant (STING ligand, cGAMP (*32*)), or two doses of mRNA-S plus an intranasal of adenovirus type 5 encoding S protein (Ad5-S) (Fig. 4A). We focused on intranasal immunization in mRNA-immunized mice, in keeping the contention that induction of mucosal immunity likely occurs in previously vaccinated individuals who will be willing to receive mucosal booster vaccines. mRNA plus Ad5-S vaccination induced greatly increased BAL RBD-specific B cells (Fig. 4B). Furthermore, mRNA plus Ad5-S vaccination induced potent mucosal CD8^+^ and CD4^+^ T cell responses but not in the spleen (Fig. S5). mRNA immunization, with or without mucosal booster immunization, induced strong circulating S1 or RBD-specific IgG in the blood and the BAL (Fig. S6 A, B). A third dose of mucosal immunization of S protein, with S trimer plus cGAMP or Ad5-S, resulted in significant increases of both S1 and RBD-specific IgA in the BAL (Fig. 4C), with Ad5-S booster inducing the highest RBD-specific IgA in the respiratory mucosa. Ad5-S booster also generated significantly higher levels of plasma IgA than other groups (Fig. S6C).

**Fig. 4.**
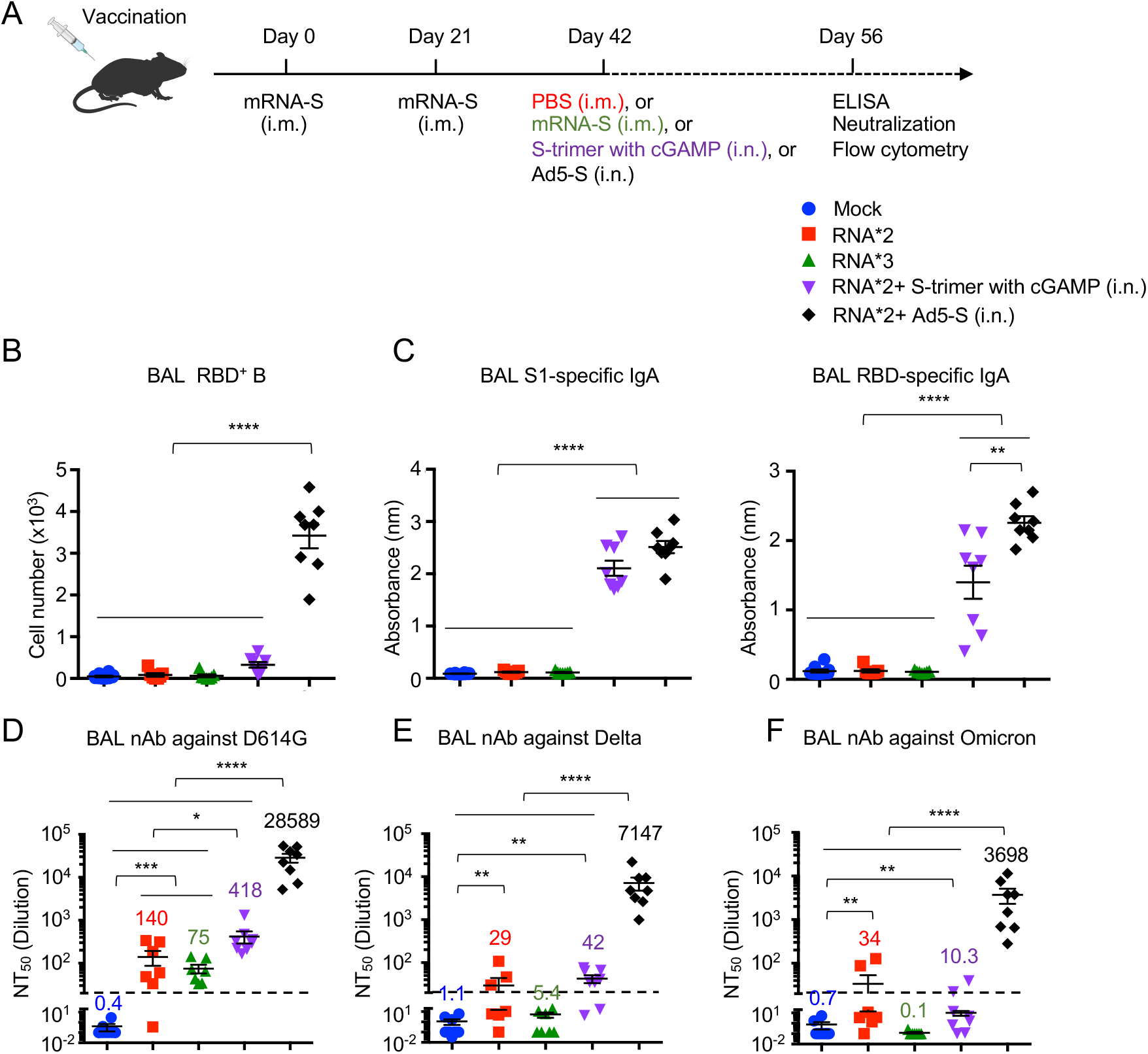
Combination of mRNA plus mucosal adenovirus immunization induces high levels of mucosal neutralizing activity against SARS-CoV-2 Omicron. C57BL/6 mice were immunized as indicated. **(A)** Schematic of vaccination strategy and experimental parameters; n=10 for control (mock), n=7 for two doses of RNA (RNA*2), n=7 for three doses of RNA (RNA*3), n=8 for two doses of RNA plus S-trimer with cGAMP booster (RNA*2+ S-trimer with cGAMP (i.n.), and n=8 for two doses of RNA plus Ad5-S booster (RNA*2+ Ad5-S (i.n.)). **(B)** Cell numbers of RBD^+^ B in the BAL following immunization. **(C)** Levels of S1- and RBD-specific IgA were measured from BAL. **(D to F)** NT_50_ of BAL against SARS-CoV-2 S D614G (D), Delta (E) and Omicron (F) pseudotyped virus were measured. nAb, neutralizing antibody. Data are means ± SEM. Statistical differences were determined by one-way ANOVA and p values were indicated by * (p < 0.05), ** (p < 0.01), *** (p < 0.001) and **** (p < 0.0001).

All immunized groups showed strong neutralization against D614G and the Delta variant in the plasma, although three-dose mRNA, or two-dose mRNA plus Ad5-S booster, vaccination induced higher levels of neutralizing Ab compared to two doses of mRNA immunization (Fig. S6 F, G). As would be expected, the mouse plasma neutralization activities against Omicron were also dramatically reduced relative to D614G or Delta (Fig. S6 H), indicating that Omicron is capable of escaping immunization-indued neutralizing Ab responses in the mouse blood similar to that in humans. However, we were still able to detect neutralizing Ab activities, at approximately similar levels, against the Omicron variant in all immunized groups (Fig. S6H).

The neutralizing Ab activity in the BAL of mRNA immunized mice (two doses or three doses) were generally lower than those in the blood, but clearly detectable against D614G, with ∼4-fold reduction in Delta, yet were around the limit of detection for the Omicron variant (Fig. 4 D-F). Strikingly, mRNA plus Ad5-S significantly increased the neutralization titer against the ancestral D614G by approximately 3 logs compared to other vaccination groups, and more importantly, maintained the strong neutralization activity against Delta as well as the Omicron variant (Fig. 5 E-F). These data indicated that compared to systemic mRNA booster, the mucosal Ad5-S booster elicits broadened Ab neutralization in the BAL against VOC. Thus, we have here identified a promising immunization strategy that can induce potent mucosal neutralizing Ab effectively against the Omicron variant.

## Discussion

mRNA vaccination elicited at least comparable neutralizing Ab levels as COVID-19 convalescents in the circulation, but generated considerably lower mucosal IgA and neutralizing Ab responses against SARS-CoV-2 D614G and Delta variants than those of convalescents, indicating that the overall magnitude of mucosal Ab responses is suboptimal following vaccination. Of note, the Omicron variant almost completely escaped the neutralization activity of BAL from either vaccinated or previously infected individuals. Additionally, we provide compelling real world evidence that mRNA vaccination does not induce notable tissue-residing S-specific memory B and T cells. Thus, despite the induction of robust circulating humoral and cellular immunity, current COVID-19 mRNA vaccines likely do not provoke sufficient levels of mucosal immunity in the human lower respiratory tract that would be needed for immediate clearance of the infectious Omicron strain to prevent the establishment of viral infection. Such a notion is consistent with the fact that Omicron continues to spread at a rapid pace in regions with high rates of vaccination and/or prior natural infection.

Our data do not dispute the notion that current vaccines are highly effective in preventing hospitalization and death. The prevention of severe disease after infection is conferred mainly by memory T and B cells (*22, 33*). To this end, CD8 T cell epitopes within Omicron Spike protein remain conserved to those of ancestral strains (*34, 35*). Thus, even though Omicron is able to breach the defense of mucosal neutralizing Ab to cause infection, the recruitment of vaccine-induced circulating memory T cells during SARS-CoV-2 breakthrough infection enables protection that restrains further viral dissemination, preventing severe disease development following infection (Fig. S7). Nevertheless, these data suggest that mucosal humoral immunity is particularly vulnerable for immune escape by Omicron. It is thus quite likely that the current vaccine strategy, even with further boosters, will not achieve “herd immunity” or prevent the occurrence of new infections or re-infections with future VOCs, particularly those with immune-evasive properties like Omicron. Thus, our findings have significant public health implications.

Our data suggest that a mucosal SARS-CoV-2 booster vaccine may be necessary to achieve more robust immunity and protection from re-infection by future variants. To this end, we have provided a proof-of-principle experiment that systemic mRNA plus mucosal Ad5-S booster provoked strong cellular immunity in the respiratory tract, and compelling mucosal IgA and neutralizing activity against Omicron. Mucosal adenovirus delivery has potential concerns regarding safety and applicability on a large scale. However, if proven to be safe, such a platform would be of great translational and clinical relevance. Alternatively, a vaccine platform with virus-like nanoparticles (*36*), which can provide strong adjuvant activity and prolonged antigen presentation *in vivo*, may also be a promising approach to boost mucosal neutralizing immunity against Omicron or future VOCs.

Our study has several limitations. Due to the highly invasive nature of the BAL procedure, we were not able to recruit a large cohort of study participants. Furthermore, the study procedure made it challenging to time recruitment or perform a longitudinal analysis; rather it enabled a snapshot of vaccination or infection-induced mucosal immunity. Additionally, most of the participants were older and may not be representative of the entire vaccinated population, although this age group is considered as the primary targeting population for vaccination as they are at highest risk of infection associated with mortality and complications. These limitations need to be addressed by future studies. Nevertheless, we provide key evidence detailing mucosal humoral and cellular immunity following vaccination in the respiratory tract. Our study highlights the importance of focusing on vaccine-induced mucosal immunity (*37*) and argues for the necessity of a mucosal booster strategy in addition to the current approach of intramuscular COVID-19 vaccines.

## Data Availability

All data produced in the present study are available upon reasonable request to the authors

## Acknowledgements

We thank the study participants for their willingness to participate in the study. This work is partially funded by the US National Institutes of Health grants AI147394, AG047156 and AG069264 to J.S.; AI147394S1 to J.S. and R.V. This work in the Liu lab was supported by a fund provided by an anonymous private donor to The Ohio State University and NCI U54CA260582 to S.-L.L. The content is solely the authors’ responsibility and does not necessarily represent the official views of the NIH. S.-L.L. was additionally supported in part by NIH R01 AI150473. M.H.K. was supported by AI057459; H.H. was supported by AI147903 and AI157852.

## Author contributions

Conceptualization: T.J., J.S. S.L.L. Acquisition of data: T.J., C.Z., T.M.C., C.L., Y.M.S., I.S.C., Y.W., S.B., D.N.S., J.P.U., J.S.R., D.E.M., J.J.M., E.S.E., R.K. Analysis and interpretation of data: T.J., C.Z., J.S., S.L.L., L.B., M.H.K., R.V. Critical reagents: J.J.T., A.J.J., M.G.A., D.W., H.H. Writing – first draft: T.J. and J.S. Funding Acquisition: J.S., S.L.L., R.V., M.H.K., H.H. and manuscript editing: S.L.L., C.Z., L.B., M.H.K., R.V.

## Methods

### Study cohorts

BAL or blood samples were collected from unvaccinated donors, vaccinated individuals, or SARS-CoV-2 infected individuals at Mayo Clinic under protocols approved by Mayo Clinic Institutional Review Boards (protocol ID 19-012187). Study participants included non-pregnant adults who were undergoing flexible bronchoscopy as part of their clinical management. However, participants who had presence of hereditary respiratory diseases (such as cystic fibrosis), clinical history of primary aspiration, neuromuscular problems, primary or secondary immune deficiencies, invasive viral or bacterial infections or a cancer diagnosis were excluded in the study. Informed consent for the use of BAL, blood and their derivatives for research was obtained from all enrolled individuals. For COVID-19 convalescents, three unvaccinated and three vaccinated samples were from a cohort that were previously recruited (*13*). Most of the vaccinated subjects received two doses of Pfizer/bioNTech (BNT162b2) or Moderna (mRNA-1273) mRNA vaccination, with three individuals receiving the third booster vaccination and one individual having the J&J (Ad26.COV2.S) vaccination. All vaccinated samples were obtained within 8 months post vaccination. Full cohort and demographic information are provided in **Extended table.1**. or have been described previously (*13*).

### BAL collection

Fiberoptic bronchoscopy and BAL were performed using moderate conscious sedation using standard clinical procedural guidelines in an outpatient bronchoscopy suite. Conscious sedation was administered in accordance with hospital policies, and a suitably trained registered nurse provided monitoring throughout the procedure. The scope was wedged into an affected segment predetermined by review of CT scan. About 100 to 200 ml of saline were instilled in 20-ml aliquots until 60 ml of lavage fluid was obtained. The specimen was placed on ice and immediately hand carried to laboratory for analysis. The fluid collected was placed on ice and transferred immediately to the laboratory for processing.

### Human single-cell purification

Plasma was isolated from whole blood by centrifuging at 1,600 rpm, room temperature (RT), for 10 min. Plasma was collected and inactivated for 30 min at 56°C, then stored at −80°C for ELISA and neutralization assay. After plasma isolation, leftover blood was mixed with phosphate-buffered saline (PBS) and then gently put over on Ficoll-Paque (Cytiva, 17144002) in a 15 mL tube. Buffy coat generated by centrifuging at 400 g for 40 min at RT was collected. For single-cell purification from BAL, BAL was filtered with a 70-μm cell strainer (Falcon) and then centrifuged at 350 g for 6 min at 4°C. Supernatant was collected and aliquots were stored at −80°C for ELISA and neutralization assay. Supernatant of BAL was further concentrated for 20x using 3 kDa Amicon Ultra-15 Centrifugal Filter Unit (Millpore Sigma, UFC900324) before use. The cells were collected for flow cytometry analysis.

### Mice vaccination and sample collection

Antigens encoded by the mRNA vaccines were derived from SARS-CoV-2 isolate Wuhan-Hu-1 (GenBank MN908947.3). Nucleoside modified mRNAs expressing SARS-CoV-2 full-length spike with two proline mutations (mRNA-S) were synthesized by in vitro transcription using T7 RNA polymerase (MegaScript, Ambion) as previously reported (*38*). mRNAs were formulated into lipid nanoparticles (LNP) using an ethanolic lipid mixture of ionizable cationic lipid and an aqueous buffer system as previously reported (*39*). Formulated mRNA-LNPs were prepared according to RNA concentrations (∼1μg/μl) and were stored at -80°C for animal immunizations. 8- to 10-week-old female mice were vaccinated with two doses of 1 μg mRNA-S with a 21-day interval. Another 21 days later, mice were boosted with PBS, 1 μg mRNA-S intramuscularly, 2 μg S-trimer (Sino Biological, 40589-V08H9) adjuvanted with 10 μg 2’3’-cGAMP (Invivogen, tlrl-nacga23) intranasally, or 10^9^ adenovirus type 5 encoding S protein (Ad5-S) (University of Iowa Viral Vector Core) intranasally after anesthetized by intraperitoneal injection of ketamine and xylazine. Three doses of PBS administered mice were used as control. 14 days later, mice were euthanatized. BAL, blood and splenocytes were collected for analysis. Isolated plasma inactivated for 30 min at 56°C and supernatant of the first 600 μL BAL collected were stored at −80°C for ELISA and neutralization assay. The cells were collected for flow cytometry analysis.

### Binding antibody response against SARS-CoV-2

General ELISA method has been previously described (*13*). Briefly, recombinant SARS-CoV-2 proteins including RBD (Sino Biological, 40592-V08H), spike S1 D614G (S1, 40591-V08H3) (Sino Biological), were precoated to 96-well plates overnight at 4°C. The following day, plates were washed with wash buffer (0.05% Tween 20 in PBS) and then blocked with Assay dilution buffer (Biolegend, 421203) for 1 hour at RT. Plasma or 20x concentrated BAL from unvaccinated donors, vaccinated and convalescents were diluted in “Assay dilution buffer” starting at a 1:5 or 1:1 dilution, respectively, and then serially diluted by 5. Plasma from mice were diluted starting at 1:100 dilution, and then serially diluted by 5. BAL from mice was not concentrated or diluted. Samples were added to the plate and incubated for 2 hours at RT. After washing three times with wash buffer, secondary antibodies diluted with “Assay dilution buffer” were added to the plate and then incubated for 1 hour at RT. Secondary antibodies including anti-human IgG (Sigma-Aldrich, A6029), anti-human IgA (Hybridoma Reagent Laboratory, HP6123), anti-human IgM (Sigma Aldrich, A6907), anti-mouse IgG (SouthernBiotech,1030-05), anti-mouse IgA (SouthernBiotech, 1040-05), anti-mouse IgM (SouthernBiotech, 1020-05) were diluted as indicated respectively. Plates were washed three times and then developed with 3,3’,5,5’ tetramethyl benzidine (TMB) buffer (Thermo Fisher Scientific, 00-4201-56) for 10 min at RT. Sulfuric acid (2 M) was used as STOP buffer. Plates were read at about 5 minutes on a microplate reader (Molecular Devices) at 450 nm with SoftMax Pro Software. The optical density (OD) value at 1:5 dilution for human plasma, 1:1 dilution for human BAL, 1:100 for mice plasma (1:500 for IgG), or original mice BAL were displayed, respectively; one dot represents each individual.

### Neutralizing antibody response against SARS-CoV-2

Virus neutralization assays were performed as previously reported (*18*). Briefly, in a 96-well format, plasma or BAL were diluted starting at a 1:40 or 1:20 dilution, respectively, and then serially diluted by a factor of 4. The viruses were incubated with plasma or BAL for 1 hour at 37°C, followed by infection of 2×10^4^ pre-seeded HEK293T-ACE2 cells on a 96-well polystyrene tissue culture plate. Gaussia luciferase activity in cell culture media was assayed 48 hours and 72 hours after infection. Note that, to ensure valid comparisons between SARS-CoV-2 variants, equivalent amounts of infectious virus were used based on the pre-determined virus titers and samples of different variants were loaded side by side in each plate. Neutralizing titer 50% (NT_50_) for each sample was determined by non-linear regression with least squares fit in GraphPad Prism 5 (GraphPad Software).

### Flow Cytometry analysis

General ELISA method has been previously described (*13*). Fresh mice and human cells or frozen human PBMC or BAL cells recovered and rested overnight in 37°C, 5% CO_2_ incubator, were washed with FACS buffer (1% FBS and 0.5 M EDTA in PBS), then stained with antibodies as listed in **Table. S2** for human and **Table. S3** for mice. Intracellular Cytokine Staining (ICS) was performed to detect vaccine-specific T cell response. Briefly, Cells were washed with FACS buffer and resuspended with complete RPMI with 10 mM HEPES supplemented with 10% FBS, 2-Mercaptoethanol, Sodium Pyruvate, Non-Essential Amino Acids, Pen-Strep, and L-Glutamine. Cells were then stimulated with 1 µg/ml S peptide pool (JPT, PM-WCPV-S) for stimulation for 6 hours (PBMC for 16 hours). In the last 4 hours of incubation, protein transport inhibitor Brefeldin-A was added. Cells stimulated with PMA/Ionomycin or DMSO only were included as positive control and negative control, respectively. Following stimulation, cells were first stained for surface markers on ice for 30 min. After washing with PBS, cells were resuspended with Zombie-dye for viability staining and incubated at room temperature (RT) for 15 min. Following surface and viability staining, cells were fixed with fixation buffer (Biolegend, 420801) and permeabilized with perm/wash buffer (Biolegend, 421002), followed by intracellular cytokine staining on ice for 30 min. Cells were then washed with perm/wash buffer and resuspended with FACS buffer. To detect RBD-specific B cells, recombinant RBD proteins, which were generated in the laboratory of Taylor lab, were incubated with the cells for 30 min at 4°C. RBD-PE and RBD-APC double-positive B cells were identified as RBD^+^ B cells. To detect S_539-546_ epitope specific CD8^+^ T cells, SARS-CoV-2 S_539-546_ MHC class I tetramer (H-2K_b_) (NIH Tetramer Core) was incubated with the cells for 30 min at 4°C. CD44^+^ Tetramer^+^ positive CD8^+^ T cells were identified as S_539-546_ epitope specific CD8^+^ T cells. Cell population data were acquired on a multi-spectral flow cytometer Cytek Aurora (Cytek Biosciences) or Attune NxT (Thermo Fisher Science) and analyzed using FlowJo Software (10.8.1, Tree Star). All human data from cytokines production assay were background-subtracted using paired DMSO treated control samples.

### Statistical analysis

Statistical tests are indicated in the corresponding figure legends. One way ANOVA was used in multi group comparison. Paired t test was used in PBMC and BAL paired comparison. Others were analyzed using independent t test. All tests were performed with a nominal significance threshold of P < 0.05, which displayed by a single asterisk. P > 0.05 was displayed by ns, means not significant. Two asterisks indicate P < 0.01, Three asterisks indicate P < 0.001, four asterisks indicate P < 0.0001.

---

**Fig. S1.**
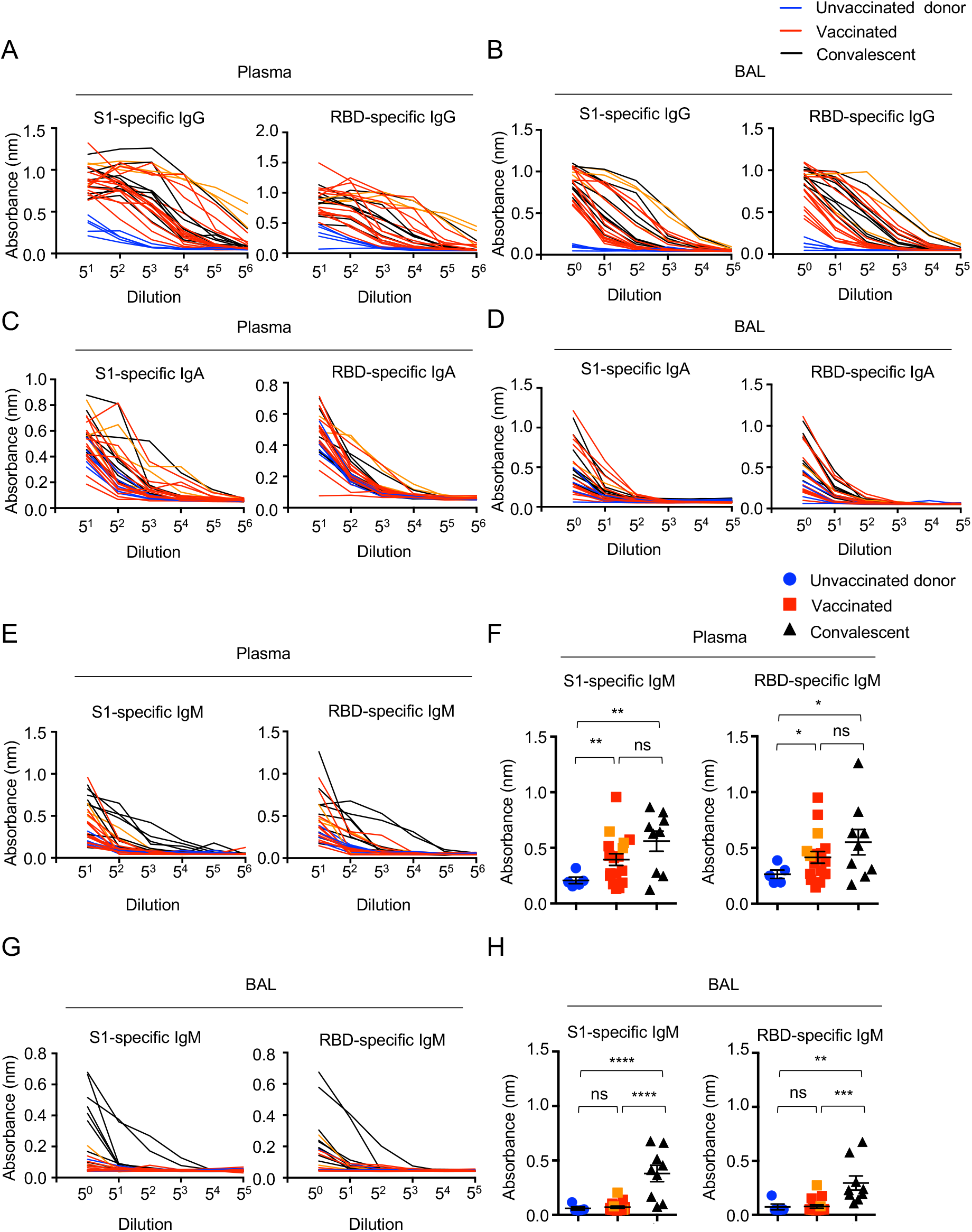
SARS-CoV-2 spike binding IgG, IgA and IgM responses in human plasma and BAL. **(A and B)** Levels of SARS-CoV-2 S1 or RBD binding IgG in the plasma (A) and bronchoalveolar fluid (BAL) (B) from unvaccinated donors, COVID-19 vaccinated or convalescents. **(C and D)** Levels of SARS-CoV-2 S1 or RBD binding IgA in the plasma (C) and bronchoalveolar fluid (BAL) (D) from unvaccinated donors, COVID-19 vaccinated or convalescents. (**E and F)** Levels of SARS-CoV-2 S1 or RBD binding IgA in the plasma from unvaccinated donors, COVID-19 vaccinated or convalescents. **(G and H)** Levels of SARS-CoV-2 S1 or RBD binding IgA in the BAL from unvaccinated donors, COVID-19 vaccinated or convalescents. Three individuals who received the booster (BNT162b2 or mRNA-1273) were indicated as orange in vaccinated group. Enrolled donors’ demographics provided in Table S1. Data in f and h are means ± SEM. Statistical differences were determined by one-way ANOVA and p values were indicated by ns, not significant (P > 0.05), * (p < 0.05), ** (p < 0.01), ***(p < 0.001) and **** (p < 0.0001).

**Fig. S2.**
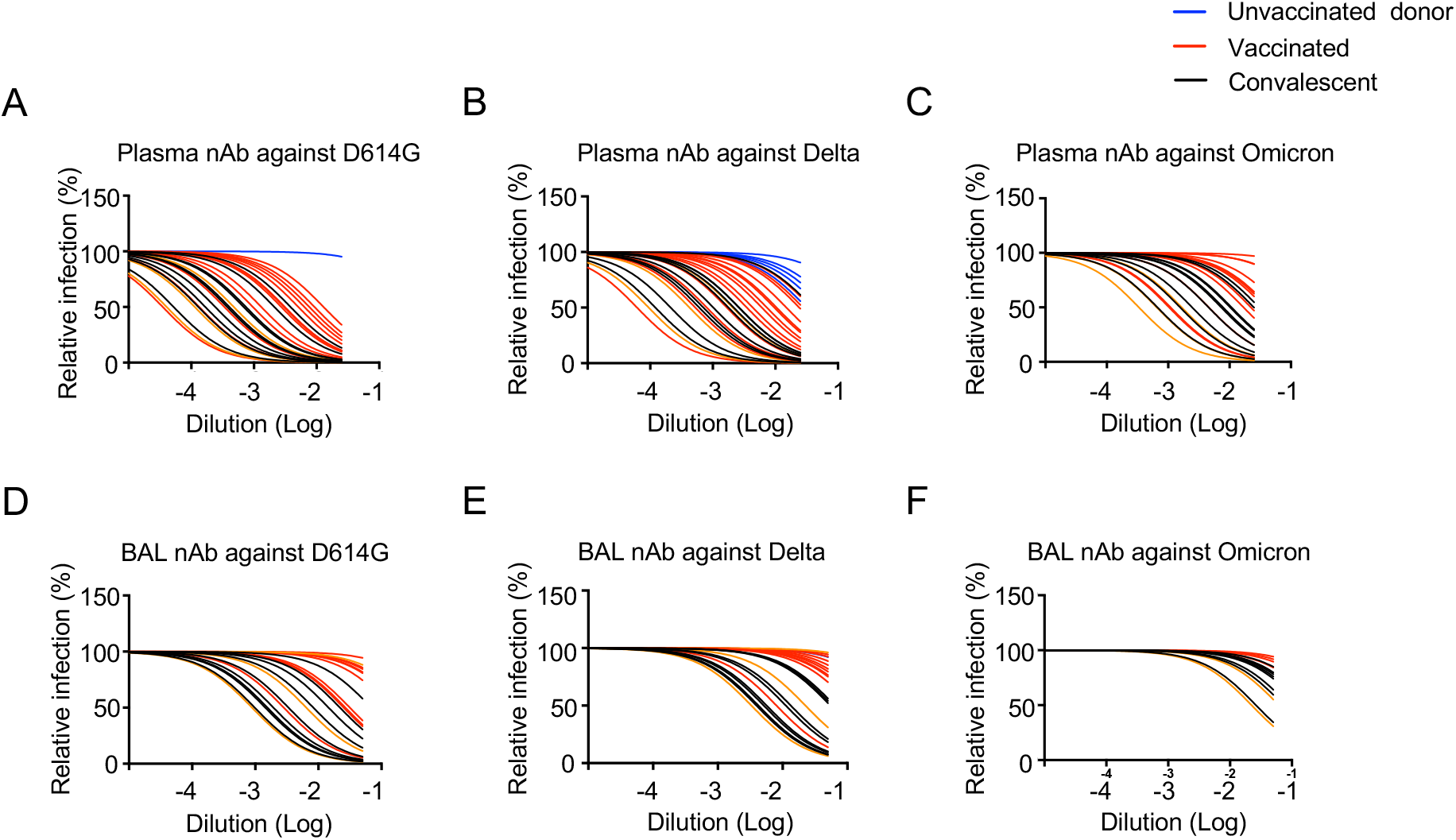
SARS-CoV-2 neutralizing antibody activities in human plasma and BAL against D614G, Delta, and Omicron. (**A to C)** Relative infection of SARS-CoV-2 S D614G (A), Delta (B) and Omicron (C) pseudotyped virus with incubation of serially diluted plasma from unvaccinated donors, vaccinated and convalescent individuals. HEK293T-ACE2 cells used as targeted cells. **(D to F)** Relative infection of SARS-CoV-2 S D614G (D), Delta (E) and Omicron (F) pseudotyped virus with incubation of serially diluted BAL from unvaccinated donors, vaccinated and convalescent individuals. HEK293T-ACE2 cells were used as targeted cells for infection. Three individuals who received the booster (BNT162b2 or mRNA-1273) were indicated as orange in vaccinated group. nAb, neutralizing antibody.

**Fig. S3.**
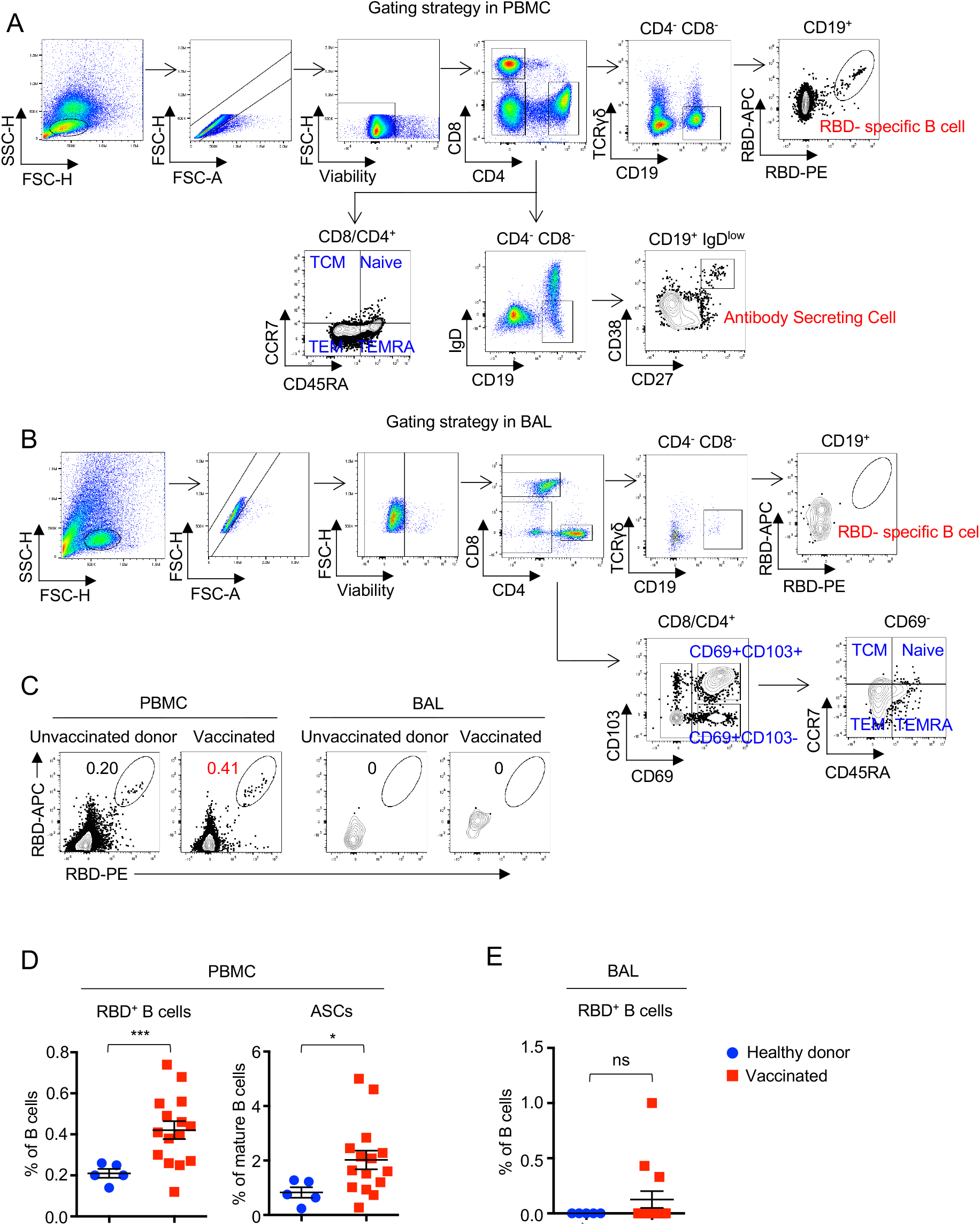
B cell response in human PBMC and BAL. **(A and B)** Gating strategy of B cells and T cells in human PBMC (A) and BAL (B). (**C)** Representative flow cytometry plots of RBD-specific B cells in the PBMC and BAL from unvaccinated donors and vaccinated. (**D)** Frequencies of RBD-specific B cells and antibody-secreting cells (ASCs) in the PBMC from unvaccinated donors and vaccinated. (**E)** Frequencies of RBD-specific B cells in the BAL from unvaccinated donors and vaccinated. Data in (D) and (E) are means ± SEM. Statistical differences were determined by independent t test and p values were indicated by ns (not significant), * (p < 0.05), and *** (p < 0.001).

**Fig. S4.**
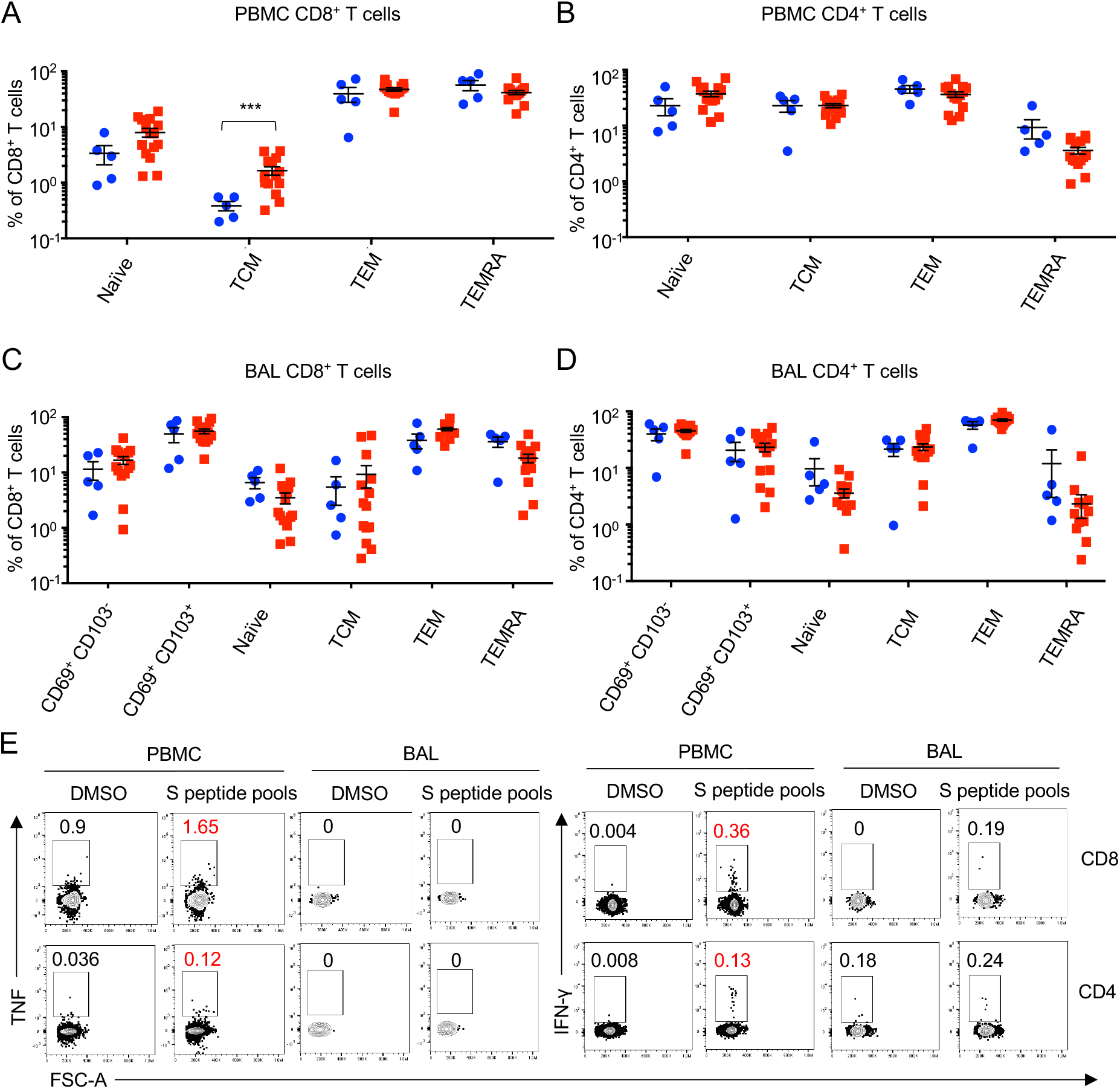
T cell response in human PBMC and BAL. **(A and B)** Frequencies of CD8^+^ (A) or CD4^+^ (B) T cell subsets in the PBMC from unvaccinated donors and vaccinated. **(C and D)** Frequencies of CD8^+^ **(C)** or CD4^+^ **(D)** T cell subsets in the BAL from unvaccinated donors and vaccinated. **(E)** Representative flow cytometry plots of TNF- and IFN-γ-producing CD8^+^ and CD4^+^ T cells in the PBMC and BAL from unvaccinated donors and vaccinated after S peptide pools stimulation. TCM, central memory T cell. TEM, effector memory T cell, TEMRA, effector memory T cell re-expressing CD45RA. Data in a is means ± SEM. Statistical differences were determined by independent t test and p values were indicated by *** (p < 0.001).

**Fig. S5.**
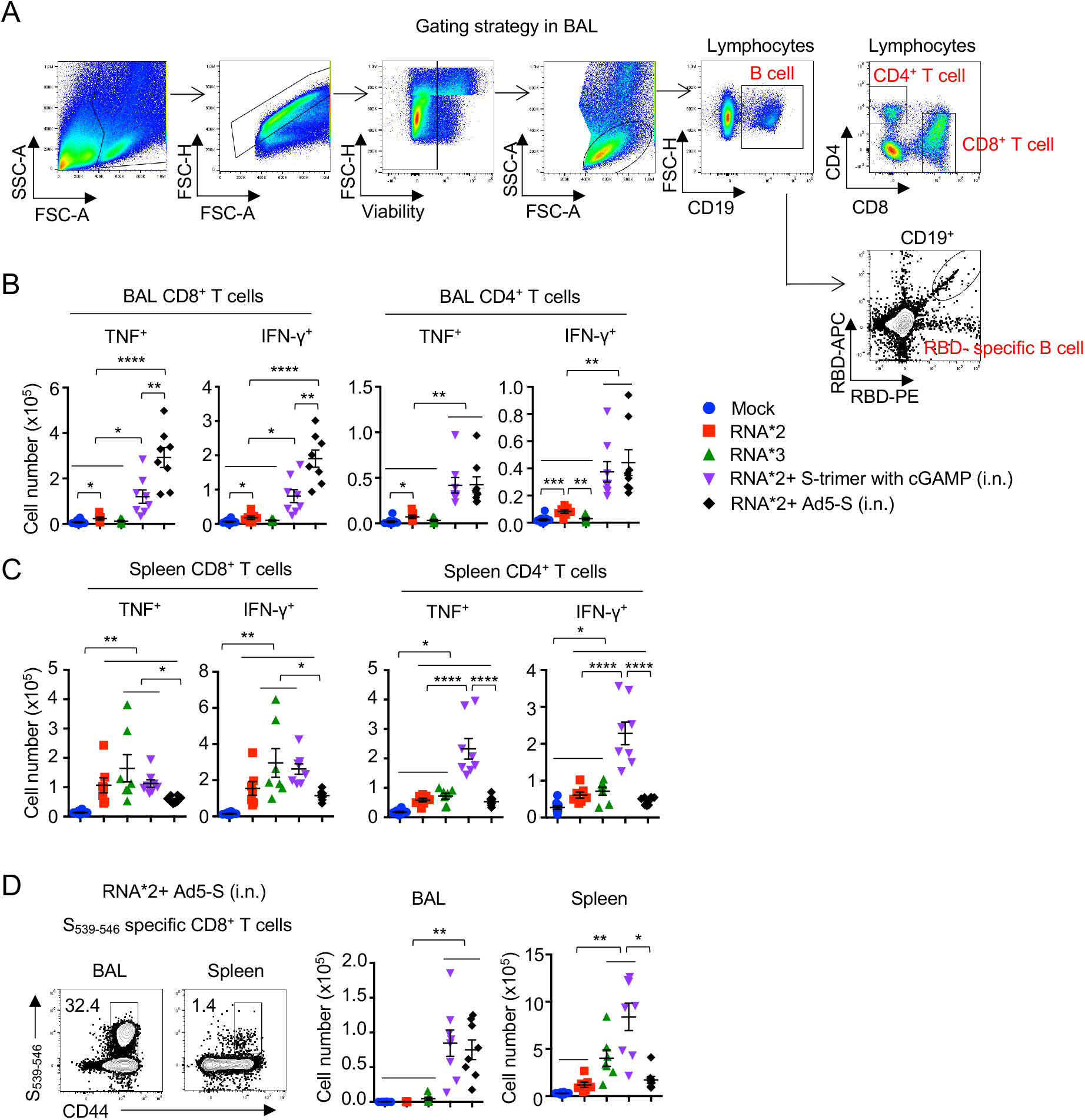
T cell responses in mouse BAL and spleen. C57BL/6 mice were immunized as indicated immunization strategies; 14 days later, T cell responses were measured from BAL and spleen. **(A)** Gating strategy of B cells and T cells in the BAL. **(B and C)** Cell numbers of cytokines producing CD8^+^ and CD4^+^ T cells in the BAL (b) and spleen (c) after S peptide pools stimulation from different groups. **(D)** Representative flow cytometry plots of CD44^+^ S_539-546_^+^ cells in CD8^+^ T cells were presented as S_539-546_ specific CD8^+^ T cells (left panel); cell numbers of S_539-546_ specific CD8^+^ T cells in the BAL and spleen from two doses of RNA plus Ad5-S booster group (right panel). Data in b-d are means ± SEM. Statistical differences were determined by one-way ANOVA and p values were indicated by * (p < 0.05), ** (p < 0.01), and **** (p < 0.0001).

**Fig. S6.**
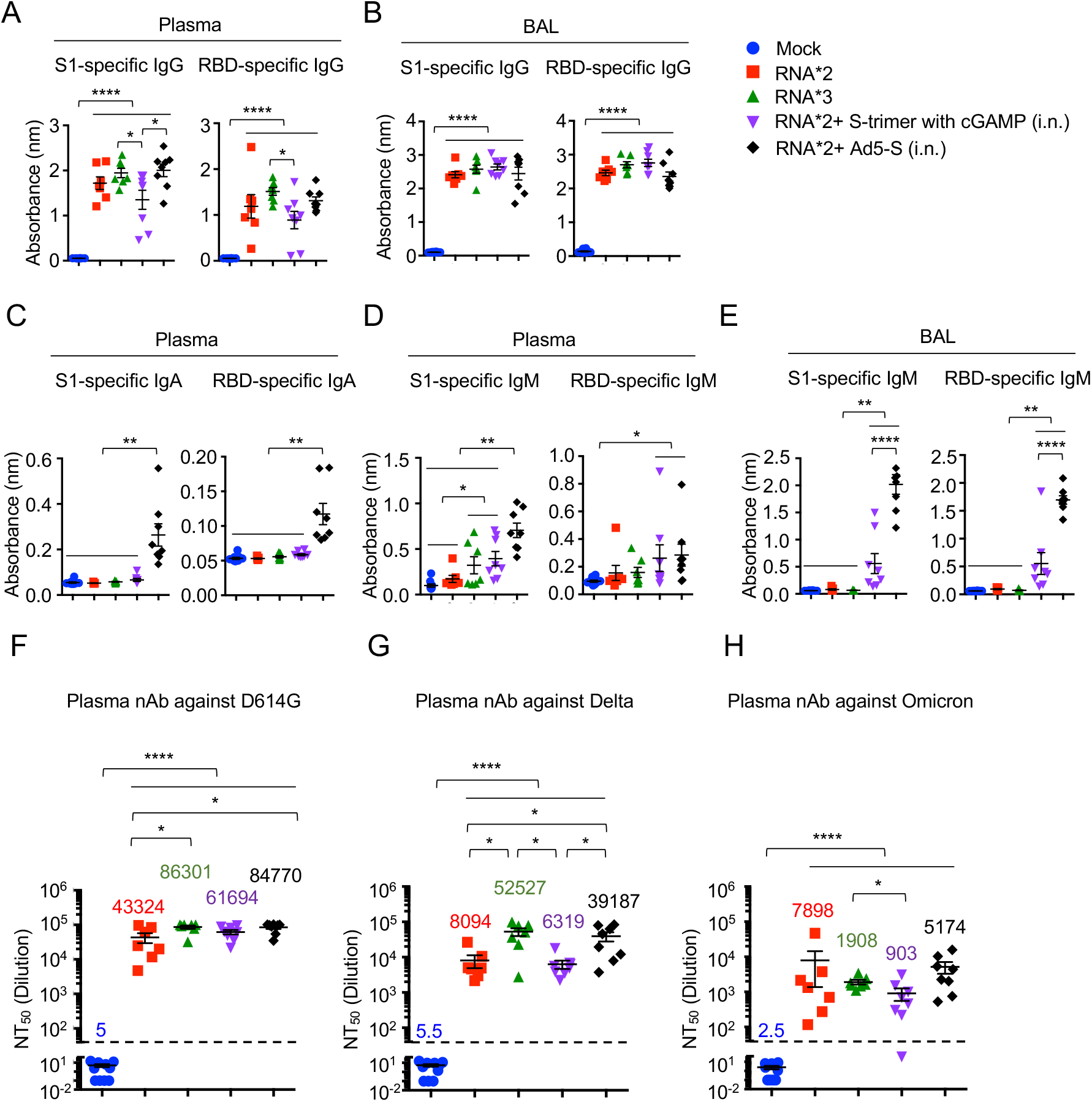
Antibody responses in mouse plasma and BAL. C57BL/6 mice were immunized as indicated immunization strategies. 14 days later, binding antibody and neutralizing antibody responses were measured from plasma and BAL. **(A and B)** Levels of S1- and RBD-specific IgG were measured from plasma (A) and BAL (B). **(C)** Levels of S1- and RBD-specific IgA were measured from plasma. **(D and E)** Levels of S1- and RBD-specific IgG were measured from plasma (D) and BAL (E). **(F to H)** NT_50_ of plasma against SARS-CoV-2 S D614G (F), Delta (G) and Omicron (H) pseudotyped virus were measured. nAb, neutralizing antibody. Data in B-D are means ± SEM. Statistical differences were determined by one-way ANOVA and p values were indicated by * (p < 0.05), ** (p < 0.01), and **** (p < 0.0001).

**Fig. S7.**
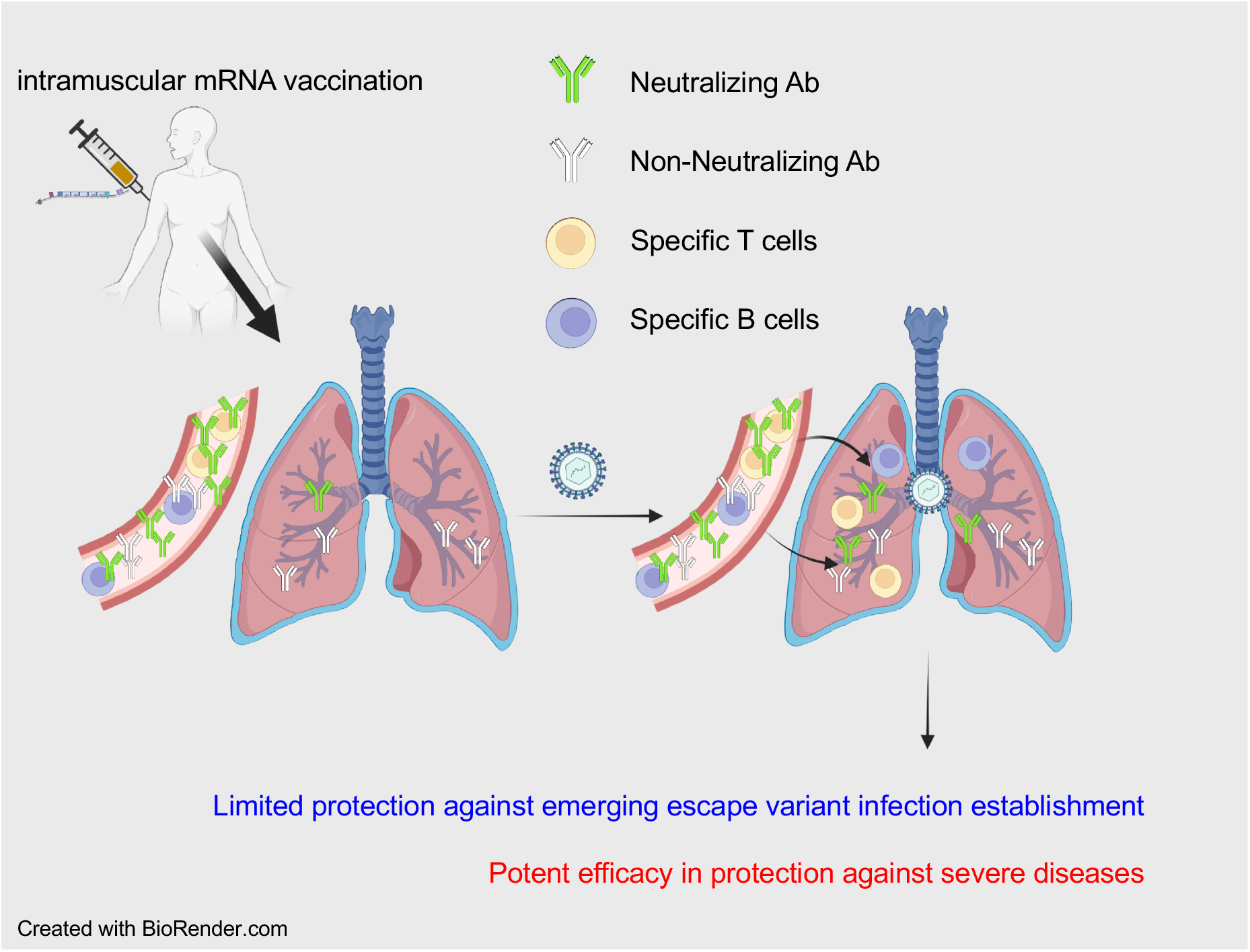
Model on the mechanism of protection by intramuscular mRNA vaccine-induced adaptive immunity against COVID-19 severe diseases but not SARS-CoV-2 infection. In this model, robust neutralizing Ab, specific T cell and B cell responses are present in circulating blood but limited in the respiratory tract post intramuscular mRNA vaccination. Adaptive immunity in local site fails to prevent respiratory infection by escape variants, but potently protects against severe diseases with the recruitment of systemic humoral and cellular immunity into the infection site upon SARS-CoV-2 escape variant infection.

**Table S1.** Enrolled donors

| ID<br>NUMBER | GENDER | AGE | VACCINE<br>RECEIVED | BOOSTER<br>RECEIVED | DAY POST<br>LAST<br>VACCINATION |
| --- | --- | --- | --- | --- | --- |
| 1 | M | 61-70 | PFIZER/<br>BIONTECH | N | 51 |
| 2 | M | 61-70 | MODERNA | N | 71 |
| 3 | M | 71-80 | MODERNA | N | 92 |
| 4 | F | 61-70 | NONE | N/A | N/A |
| 5 | F | 71-80 | NONE | N/A | N/A |
| 6 | F | 71-80 | MODERNA | N | 87 |
| 7 | F | 61-70 | PFIZER/<br>BIONTECH | N | 124 |
| 8 | M | 71-80 | JANSSEN<br>(J&J) | N | 107 |
| 9 | M | 31-40 | PFIZER/<br>BIONTECH | N | 95 |
| 10 | M | 61-70 | PFIZER/<br>BIONTECH | N | 104 |
| 11 | F | 81-90 | MODERNA | N | 187 |
| 12 | F | 71-80 | MODERNA | N | 206 |
| 13 | F | 71-80 | MODERNA | N | 165 |
| 14 | M | 61-70 | PFIZER/<br>BIONTECH | N | 171 |
| 15 | 71-80 | 71-80 | MODERNA | N | 235 |
| 16 | F | 41-50 | PFIZER/<br>BIONTECH | N | 206 |
| 17 | M | 71-80 | MODERNA | Y | 34 |
| 18 | M | 61-70 | PFIZER/<br>BIONTECH | Y | 57 |
| 19 | M | 61-70 | PFIZER/<br>BIONTECH | Not sure | < 165 |
| 20 | M | 61-70 | MODERNA | Y | 51 |

**Table S2.** List of human antibody used for flow cytometry

| ANTIGEN | FLUORESCENCE | CLONE | COMPANY |
| --- | --- | --- | --- |
| CD103 | APC/Fire750 | Ber-ACT8 | Biolegend |
| CD19 | BV786 | SJ25C1 | BD Biosciences |
| CD27 | Pacific Blue | M-T271 | Biolegend |
| CD38 | BV480 | HIT2 | BD Biosciences |
| CD4 | AF532 | SK3 | ThermoFisher Scientifics |
| CD4 | BV421 | SK3 | ThermoFisher Scientifics |
| CD45RA | BV650 | HI100 | Biolegend |
| CD69 | APC-Cy7 | FN50 | Biolegend |
| CD69 | PE-eFluor610 | FN50 | ThermoFisher Scientifics |
| CD8 | PerCP-eFluor710 | OKT8 | ThermoFisher Scientifics |
| CD8 | FITC | OKT8 | ThermoFisher Scientifics |
| CCR7 | BV711 | G043H7 | Biolegend |
| IFN-γ | PE | B27 | BD Biosciences |
| IgD | BV510 | IA6-2 | Biolegend |
| TCR$\gamma\delta$ | BV750 | 11F2 | BD Biosciences |
| TNF-α | AF700 | MAB11 | ThermoFisher Scientifics |
| Viability | Zombie Aqua |  | Biolegend |
| Viability | Zombie NIR |  | Biolegend |

**Table S3.**
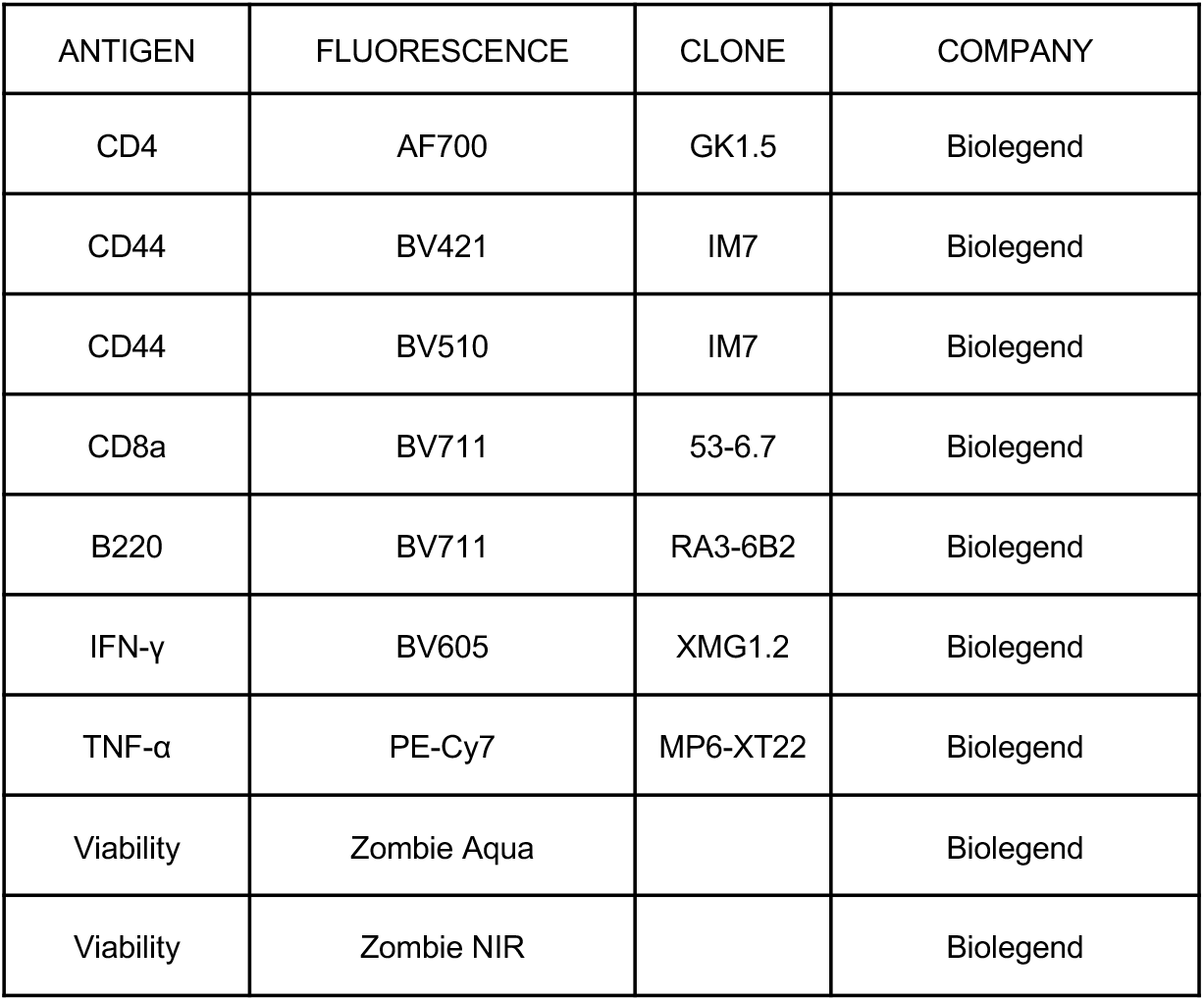
List of mouse antibody used for flow cytometry

| ANTIGEN | FLUORESCENCE | CLONE | COMPANY |
| --- | --- | --- | --- |
| CD4 | AF700 | GK1.5 | Biolegend |
| CD44 | BV421 | IM7 | Biolegend |
| CD44 | BV510 | IM7 | Biolegend |
| CD8a | BV711 | 53-6.7 | Biolegend |
| B220 | BV711 | RA3-6B2 | Biolegend |
| IFN-γ | BV605 | XMG1.2 | Biolegend |
| TNF-α | PE-Cy7 | MP6-XT22 | Biolegend |
| Viability | Zombie Aqua |  | Biolegend |
| Viability | Zombie NIR |  | Biolegend |

